# Estimation of airborne viral emission: quanta emission rate of SARS-CoV-2 for infection risk assessment

**DOI:** 10.1101/2020.04.12.20062828

**Authors:** G. Buonanno, L. Stabile, L. Morawska

## Abstract

Airborne transmission is a pathway of contagion that is still not sufficiently investigated despite the evidence in the scientific literature of the role it can play in the context of an epidemic. While the medical research area dedicates efforts to find cures and remedies to counteract the effects of a virus, the engineering area is involved in providing risk assessments in indoor environments by simulating the airborne transmission of the virus during an epidemic. To this end, virus air emission data are needed. Unfortunately, this information is usually available only after the outbreak, based on specific reverse engineering cases. In this work, a novel approach to estimate the viral load emitted by a contagious subject on the basis of the viral load in the mouth, the type of respiratory activity (e.g. breathing, speaking), respiratory physiological parameters (e.g. inhalation rate), and activity level (e.g. resting, standing, light exercise) is proposed. The estimates of the proposed approach are in good agreement with values of viral loads of well-known diseases from the literature. The quanta emission rates of an asymptomatic SARS-CoV-2 infected subject, with a viral load in the mouth of 10^8^ copies mL^−1^, were 10.5 quanta h^−1^ and 320 quanta h^−1^ for breathing and speaking respiratory activities, respectively, at rest. In the case of light activity, the values would increase to 33.9 quanta h^−1^ and 1.03×10^3^ quanta h^−1^, respectively.

The findings in terms of quanta emission rates were then adopted in infection risk models to demonstrate its application by evaluating the number of people infected by an asymptomatic SARS-CoV-2 subject in Italian indoor microenvironments before and after the introduction of virus containment measures. The results obtained from the simulations clearly highlight that a key role is played by proper ventilation in containment of the virus in indoor environments.

## 1. Introduction

Expiratory human activities generate droplets, which can also carry viruses, through the atomization processes occurring in the respiratory tract when sufficiently high speeds are reached (Chao et al., 2009; Morawska, 2006). Indeed, during breathing, coughing, sneezing or laughing, toques of liquid originating from different areas of the upper respiratory tract are drawn out from the surface, pulled thin, and broken into columns of droplets of different sizes (Hickey and Mansour, 2019). The content of infectious agents expelled by an infected person depends, among other factors, on the location within the respiratory tract from which the droplets originated. In particular, air velocities high enough for atomization are produced when the exhaled air is forced out through some parts of the respiratory tract which have been greatly narrowed. The front of the mouth is the site of narrowing and the most important site for atomization; since most droplets originate at the front of the mouth, the concentration of an infectious agent in the mouth (sputum) is representative of the concentration in the droplets emitted during the expiratory activities (Morawska, 2006). Thus, knowledge of the size and origin of droplets is important to understand transport of the virus via the aerosol route. Contrary to the findings of early investigations (Duguid, 1945; Jennison, 1942; Wells, 1934), subsequent studies involving optical particle detection techniques capable of measurements down to fractions of a micrometer suggested that the majority of these particles are in the sub-micrometer size range (Papineni and Rosenthal, 1997). More recently, the growing availability of higher temporal and spatial visualization methods using high-speed cameras (Tang et al., 2011), particle image velocimetry (Chao et al., 2009) and, above all, increasingly accurate particle counters (Morawska et al., 2009) allowed the detailed characterization and quantitation of droplets expelled during various forms of human respiratory exhalation flows (e.g. breathing, whispering, speaking, coughing). Therefore, in recent years a marked development has occurred both in the techniques for detecting the viral load in the mouth and in the engineering area of the numerical simulation of airborne transmission of the viral load emitted.

However, the problem of estimating the viral load emitted, which is fundamental for the simulation of airborne transmission, has not yet been solved. This is a missing “transfer function” that would allow the virology area, concerned with the viral load values in the mouth, to be connected with the aerosol science and engineering areas, concerned with the spread and mitigation of contagious particles.

A novel approach is here presented for estimating the viral load emitted by an infected individual. This approach, based on the principle of conservation of mass, represents a tool to connect the medical area, concerned with the concentration of the virus in the mouth, to the engineering area, dedicated to the simulation of the virus dispersion in the environment. On the basis of the proposed approach, the quanta emission rate data of SARS-CoV-2 were calculated as a function of different respiratory activities, respiratory parameters, and activity levels.

The quanta emission rate data, starting from the recently documented viral load in sputum (expressed in copies mL^−1^), were then applied in an acknowledged infection risk model to investigate the effectiveness of the containment measures implemented by the Italian government to reduce the spread of SARS-CoV-2. In particular, airborne transmission of SARS-CoV-2 by an asymptomatic subject within pharmacies, supermarkets, restaurants, banks, and post offices were simulated, and the reduction in the average number of infected people from one contagious person, R_0_, was estimated.

## 2. Materials and methods

### 2.1. Estimation of the quanta emission rate

The approach proposed in the present work is based on the hypothesis that the droplets emitted by the infected subject have the same viral load as the sputum. Therefore, if the concentration of the virus in the sputum and the quantity of droplets emitted with dimensions less than 10 µm is known, the viral load emitted can be determined through a mass balance. In particular, the viral load emitted, expressed in terms of quanta emission rate (ER_q_, quanta h^−1^), was evaluated as:

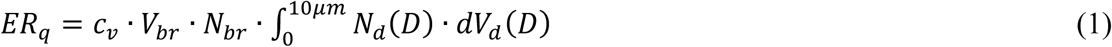

where *c*_v_ is the viral load in the sputum (RNA copies mL^−1^), *V*_br_ is the volume of exhaled air per breath (cm^3^; also known as tidal volume), *N*_br_ is the breathing rate (breath h^−1^), *N*_d_ is the droplet number concentration (part. cm^−3^), and *V*_d_(*D*) is the volume of a single droplet (mL) as a function of the droplet diameter (*D*). Information about the viral load in terms of quanta is essential as the quantum represents the “viral load” considered in engineering science: in other words, an infected individual constantly generates a number of infectious quanta over time, where a “quantum” is defined as the dose of airborne droplet nuclei required to cause infection in 63% of susceptible persons.

The volume of the droplet (*V*_d_) was determined on the basis of data obtained experimentally by (Morawska et al., 2009): they measured the size distribution of droplets for different expiratory activities (e.g. breathing, whispering, counting, speaking), recognizing that such droplets present one or more modes occurring at different concentrations. In particular, in the study a particle size distribution with four channels was considered with midpoint diameters of *D*_1_=0.8, *D*_2_=1.8, *D*_3_=3.5, and *D*_4_=5.5 µm. As an example, speaking was recognized as producing additional particles in modes near 3.5 and 5.5 µm. These two modes became even more pronounced during sustained vocalization. Details of the aerosol concentrations at the four channels of the size distribution during each expiratory activity are reported in Table 1. The midpoint diameters of each channel were used to calculate the corresponding volume of the droplets.

**Table 1.** Droplet concentrations (Ni, part. cm^−3^) of the different size distribution channels during each expiratory activity measured by (Morawska et al., 2009)

| Expiratory activity | $D_1$ (0.80 $\mu\text{m}$) | $D_2$ (1.8 $\mu\text{m}$) | $D_3$ (3.5 $\mu\text{m}$) | $D_4$ (5.5 $\mu\text{m}$) |
| --- | --- | --- | --- | --- |
| Whispered counting | 0.236 | 0.068 | 0.007 | 0.011 |
| Voiced counting | 0.110 | 0.014 | 0.004 | 0.002 |
| Speaking | 0.751 | 0.139 | 0.139 | 0.059 |
| Breathing | 0.084 | 0.009 | 0.003 | 0.002 |

Based on the results obtained by (Morawska et al., 2009), equation (1) can be simplified as:

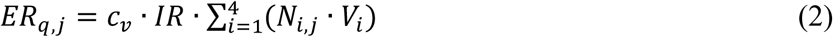

where *j* indicates the different expiratory activities considered (namely whispered counting, voiced counting, speaking, breathing) and *IR* (m^3^ h^−1^) is the inhalation rate, i.e. the product of breathing rate (*N*_br_) and tidal volume (*V*_br_), which is a function of the activity level of the infected subject. The quanta emission rate from equation (2) can vary in a wide range depending on the virus concentration in the mouth, the activity level, and the different types of expiration. Regarding the inhalation rate effect, the quanta emission rate calculations are shown for three different activity levels (resting, standing, and light exercise) in which the inhalation rates, averaged between males and females, are equal to 0.36, 0.54, and 1.16 m^3^ h^−1^, respectively (Adams, 1993; International Commission on Radiological Protection, 1994).

### 2.2. A demonstration application: the containment measures for the spread of SARS-CoV-2 in Italy

The pandemic of a novel human coronavirus, now named Severe Acute Respiratory Syndrome CoronaVirus 2 (SARS-CoV-2 throughout this manuscript), emerged in Wuhan (China) in late 2019 and then spread rapidly in the world (https://www.who.int/emergencies/diseases/novel-coronavirus-2019). In Italy, an outbreak of SARS-CoV-2 infections was detected starting from 16 cases confirmed in Lombardy (a northern region of Italy) on 21 February. The Italian government has issued government a decree dated 11 March 2020 concerning urgent measures to contain the contagion throughout the country. This decree regulated the lockdown of the country to counteract and contain the spread of the SARS-CoV-2 virus by suspending retail commercial activities, with the exception of the sale of food and basic necessities. It represents the starting point of a system with imposed constraints. Among the measures adopted for the containment of the virus in Italy, great importance was placed on the safe distance of 1 m (also known as “droplet distance”). This distance was actually indicated by the World Health Organization as sufficient to avoid transmission by air, without any reference to the possibility of transmission over greater distances indoors (https://www.who.int/emergencies/diseases/novel-coronavirus-2019). With this measure, along with the opening of only primary commercial establishments (such as pharmacies, supermarkets, banks, post offices) and the closure of restaurants, the Italian government has adopted the concept of spacing (known as “social distancing”) to prevent the spread of the infection. Obviously, this limit per se would have no influence on the reduction of airborne transmission of the infection in indoor environments since this distance is compatible with the normal gathering of people in commercial establishments. Actually, on an absolutely voluntary basis, and despite the continuous denials by the government on the risk of indoor airborne transmission, commercial associations have changed the methods of accessing their commercial spaces such as restaurants, pharmacies, supermarkets, post offices, and banks; for example, by forcing customers to queue outside. It is clear that the best choice in containing an epidemic is a total quarantine which, however, appears to have enormous costs and social impacts, especially in Western countries.

To show the possible effect of the measures imposed by the Italian government (i.e. lockdown), the infection risk in different indoor microenvironments for the exposed population due to the presence of one contagious individual was simulated, adopting the infection risk model described in section 2.2.1. In particular, the risk expressed in terms of basic reproduction number (R_0_) was derived from the quanta concentration and the infection risk; indeed, R_0_ represents the average number of secondary infections produced by a typical case of an infection in a population where everyone is susceptible (Rothman et al., 2008).

The indoor microenvironments considered here were a pharmacy, supermarket, restaurant, post office, and bank whose dimensions are summarized in Table 2. Two different exposure scenarios were simulated for each microenvironment: before lockdown (B) and after lockdown (A). In the simulation of the scenario before lockdown, the microenvironments were run with no particular recommendations; thus, people enter the microenvironments and queue indoors, often resulting in overcrowded environments. Since most of the indoor microenvironments in Italy are not equipped with mechanical ventilation systems, the simulations were performed considering two different situations: natural ventilation (a typical value for an Italian building equal to 0.2 h^−1^ was adopted, with reference to (d’Ambrosio Alfano et al., 2012; Stabile et al., 2017)) and mechanical ventilation (calculated according the national standard, UNI 10339 (UNI, 1995), as a function of the crowding index and the type of indoor microenvironment). The scenario after lockdown was tested considering the typical solutions adopted (on a voluntary basis) by the owners of stores and offices – reduced personnel, a reduced number of customers inside the microenvironment, customers forced to queue outdoors, and doors kept open. The scenario after lockdown was also tested for both natural ventilation and mechanical ventilation; in this case a slight increase in the air exchange rate (AER) for natural ventilation (0.5 h^−1^) was considered in order to take into account that the door was always kept open. The restaurant was not tested in the scenario after lockdown since such commercial activity was closed down as a consequence of the lockdown. For all the scenarios considered in the simulations, the infected individual was considered to enter the microenvironment as the first customer (alone or along with other individuals according to the scenarios summarized in Table 2). All the scenarios were simulated taking into account that the virus is able to remain viable in the air for up to 3 hours post aerosolization as recently detected by (van Doremalen et al., 2020); thus, if the infected individual remained inside the environment for 10 minutes (e.g. pharmacy), the calculation of the quanta concentration, infection risk, and R_0_ was performed for up to 3 hours and 10 minutes (named “total exposure time” in Table 2). For restaurants the calculation was performed for 3 hours considering that after 3 hours (i.e. two groups remaining inside for 1 hour and 30 minutes one after the other) the microenvironment was left empty.

**Table 2.** Summary of the exposure scenarios tested for the different microenvironments under investigation: dimensions, ventilation conditions, number of workers and customers.

|  |  | Pharmacy | Supermarket | Restaurant | Post office | Bank |
| --- | --- | --- | --- | --- | --- | --- |
| Dimensions | Floor area (A, m<sup>2</sup>) | 25 | 600 | 100 | 100 | 50 |
|  | Height (h, m<sup>2</sup>) | 3 | 3 | 3 | 3 | 3 |
|  | Volume (V, m<sup>3</sup>) | 75 | 1800 | 300 | 300 | 150 |
| Exposure scenario before lockdown (B) | Number of workers | 5 (always present) | 10 (always present) | 4 (just the waiters, always present) | 8 (always present) | 4 (always present) |
|  | Number and activity of the costumers | – 1 new customer per min entering the pharmacy, | – 1 new customer every 30 s entering the supermarket, | – 80 costumers every 1.5 hours, | – 1 new customer every 30 s entering the post office, | – 1 new customer per min entering the bank, |
|  |  | – every customer remains 10 min inside (including waiting time), | – every customer remains 30 min inside, | – restaurant working for 3 hours (evening), | – every customer remains 15 min inside (including waiting time), | – every customer remains 15 min inside (including waiting time), |
|  |  | – thus, 10 customers are simultaneously present | – thus, 60 customers are simultaneously present | – thus, 80 customers are simultaneously present for a total number of 160 customers per evening. | – thus, 30 customers are simultaneously present | – thus, 15 customers are simultaneously present |
|  | Air exchange rate (AER, h<sup>-1</sup>) for natural ventilation (NV) | 0.2 | 0.2 | 0.2 | 0.2 | 0.2 |
|  | Air exchange rate (AER, h<sup>-1</sup>) for mechanical ventilation (MV) | 2.2 | 1.1 | 9.6 | 2.4 | 2.4 |
|  | Total exposure time | 3 hours and 10 minutes | 3 hours and 30 minutes | 3 hours | 3 hours and 15 minutes | 3 hours and 15 minutes |
| Exposure scenario after lockdown (A) | Number of workers | 3 (always present) | 10 (always present) | - | 4 (always present) | 4 (always present) |
|  | Number and activity of the costumers | – 2 new customers every five min entering the pharmacy, | – 1 new customer per min entering the supermarket, |  | – 4 new customers every five min entering the post office, | – 4 new customers every five min entering the bank, |
|  |  | – every customer remains 5 min inside, | – every customer remains 10 min inside, | - | – every customer remains 10 min inside, | – every customer remains 10 min inside, |
|  |  | – people forced to queue outside the pharmacy, | – people forced to queue outside the supermarket, |  | – people forced to queue outside the post office, | – people forced to queue outside the bank, |
|  |  | – thus, 2 customers are simultaneously present | – thus, 10 customers are simultaneously present |  | – thus, 4 customers are simultaneously present | – thus, 4 customers are simultaneously present |
|  | Air exchange rate (AER, h<sup>-1</sup>) for natural ventilation (NV) | 0.5 | 0.2 | - | 0.5 | 0.5 |
|  | Air exchange rate (AER, h<sup>-1</sup>) for mechanical ventilation (MV) | 2.2 | 1.1 | - | 2.4 | 2.4 |
|  | Total exposure time | 3 hours and 5 minutes | 3 hours and 10 minutes | 3 hours | 3 hours and 10 minutes | 3 hours and 10 minutes |

#### 2.2.1. The infection risk model

The simulation of airborne transmission of SARS-CoV-2 was performed adopting the infection risk assessment typically implemented to evaluate the transmission dynamics of infectious diseases and to predict the risk of these diseases to the public. The model considered here to quantify the airborne transmitted infection risk was carried out by Gammaitoni and Nucci (Gammaitoni and Nucci, 1997) which represents an upgrade of an earlier model provided by Wells-Riley (Riley et al., 1978). This model was successfully adopted in previous papers estimating the infection risk due to other diseases (e.g. influenza, SARS, tuberculosis, rhinovirus) in different indoor microenvironments such as airplanes (Wagner et al., 2009), cars (Knibbs et al., 2011), and hospitals. The Gammaitoni and Nucci model is based on the rate of change in quanta levels through time; in particular, the differential equations for the change of quanta in a control volume as well as the initial conditions (here not reported for the sake of brevity) allowed to evaluate the quanta concentration in an indoor environment at the time *t, n*(*t*), as:

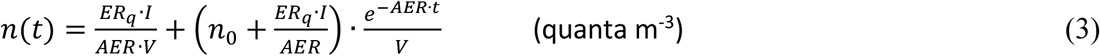

where AER (h^−1^) represents the air exchange rate of the space investigated, *n*_0_ represents the initial number of quanta in the space, *I* is the number of infectious subjects, *V* is the volume of the indoor environment considered, and ER_q_ is the abovementioned quanta emission rate (quanta h^−1^) characteristic of the specific disease/virus under investigation.

The equation was derived considering the following simplifying assumptions: the quanta emission rate is considered to be constant, the latent period of the disease is longer than the time scale of the model, and the droplets are instantaneously and evenly distributed in the room (Gammaitoni and Nucci, 1997). The latter represents a key assumption for the application of the model as it considers that the air is well-mixed within the modelled space. The authors highlight that in epidemic modeling, where the target is the spread of the disease in the community, it is impossible to specify the geometries, the ventilation, and the locations of the infectious sources in each microenvironment. Therefore, adopting the well-mixed assumption is generally more reasonable than hypothesizing about specific environments and scenarios because the results must be interpreted on a statistical basis (Sze To and Chao, 2010).

To determine the infection risk (*R*, %) as a function of the exposure time (*t*) of susceptible people, the quanta concentration was integrated over time through the Wells–Riley equation (Riley et al., 1978) as:

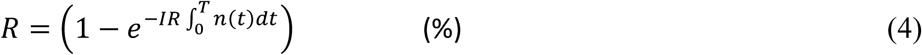

where *IR* is the inhalation rate of the exposed subject (which is, once again, affected by the subject’s activity level) and *T* is the total time of exposure (h). From the infection risk *R*, the number of susceptible people infected after the exposure time can be easily determined by multiplying it by the number of exposed individuals. In fact, equations (3) and (4) were adopted to evaluate the infection risk of different exposure scenarios of Italian microenvironments hereinafter reported. The quanta emission rate used in the simulation of the scenario represents the average value obtained from the four expiratory activities (whispered counting, voiced counting, speaking, and breathing); the data are reported and discussed in the result sections.

## 3. Results and discussions

### 3.1. The quanta emission rate

As discussed in the Materials and methods section, the quanta emission rate, ER_q_, depends on several parameters. In Figure 1 the ER_q_ (quanta h^−1^) trends are reported as a function of the viral load in the sputum (c_v_, RNA copies mL^−1^) for different expiratory activities (whispered counting, voiced counting, speaking, breathing) and different activity levels (resting, standing, light exercise). To represent the large variabilities (over several orders of magnitude) of ER_q_ as a function of c_v_, the graph is reported on a bi-logarithmic scale.

**Figure 1.**
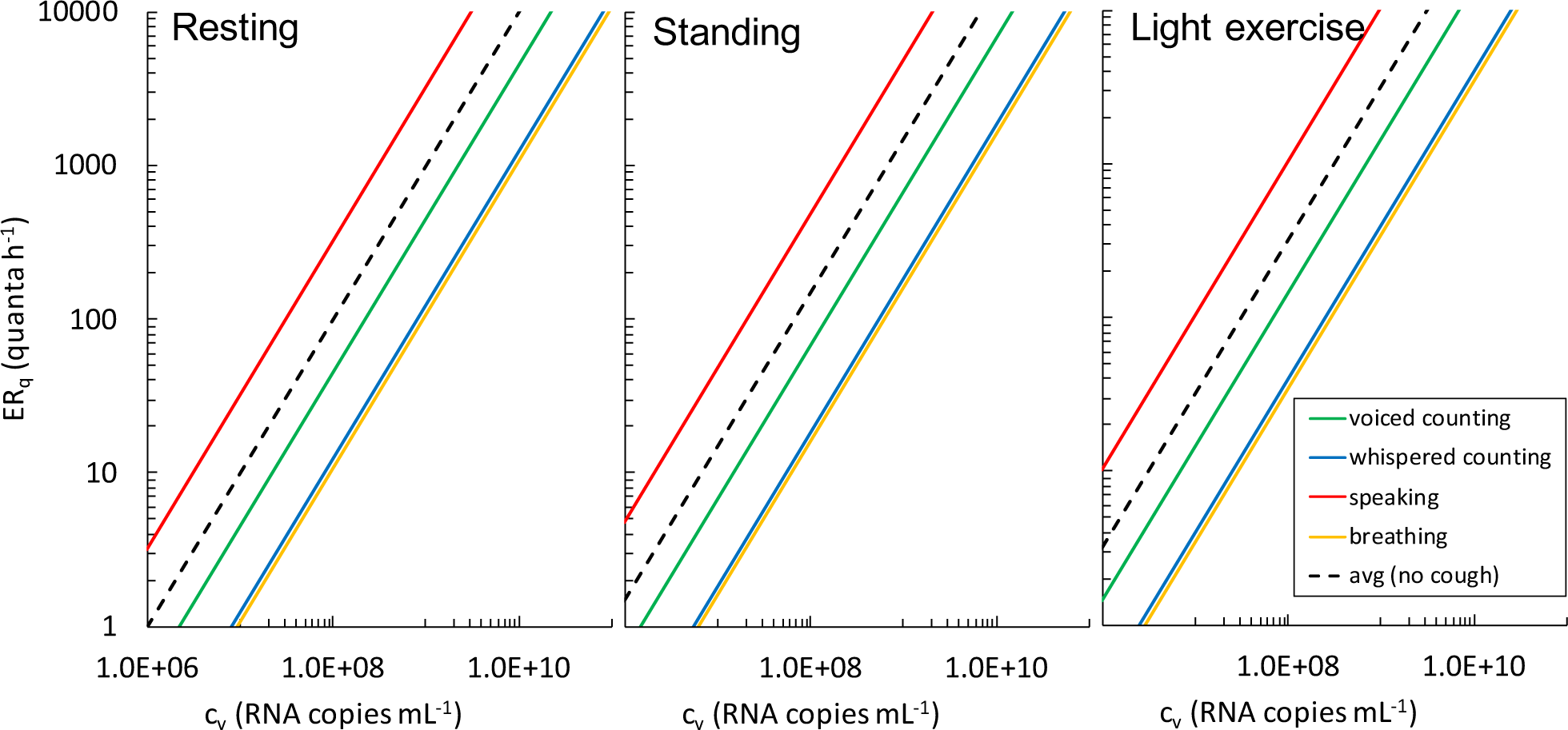
ERq (quanta h^−1^) trends as a function of the viral load in sputum (cv, RNA copies mL^−1^) for different respiratory activities (whispered counting, voiced counting, speaking, breathing) and different activity levels (resting, standing, light exercise). The average (avg) value of the respiratory activities considered was also reported.

To benchmark the proposed approach for the estimation of the quanta emission rate, we considered the case of seasonal influenza for which more data are available in terms of both viral load in sputum and quanta emission rate. As an example, (Hirose et al., 2016) found an average value of RNA concentration in sputum for influenza equal to 2.38×10^7^ copies mL^−1^. Thus, applying the findings of the proposed approach in the case of a standing subject, a corresponding ER_q_ varying between 3.7 (breathing) and 114 quanta h^−1^ (speaking) is estimated: this value is in good agreement with the quanta emission rates for influenza found in the scientific literature, from 2 to 128 quanta h^−1^ with a most frequent value of 67 quanta h^−1^ (Knibbs et al., 2012). Such variability in the quanta emission rates for influenza is due both to the method used to calculate it (Rudnick and Milton, 2003) and, especially, the viral load of the subject and the type of respiratory activity, which is typically not reported and discussed.

With reference to SARS-CoV-2 infection, researchers have recently found values for the viral load in the mouth between 10^2^ and 10^11^ copies mL^−1^, also variable in the same patient during the course of the disease (Pan et al., 2020; To et al., 2020; Woelfel et al., 2020). (Rothe et al., 2020) reported a case of SARS-CoV-2 infection acquired outside Asia in which transmission appears to have occurred during the incubation period in the index patient. A high viral load of 10^8^ copies mL^−1^ was found, confirming that asymptomatic persons are potential sources of SARS-CoV-2 infection; this may warrant a reassessment of the transmission dynamics of the current outbreak. Table 3 lists the quanta emission rates (ER_q_) for a SARS-CoV-2 infected asymptomatic subject as a function of activity level (resting, standing, and light exercise) and respiratory activity (voiced counting, whispered counting, speaking, breathing). The data confirm the huge variations in the quanta emission rate, with the lowest value being for breathing during resting activity (10.5 quanta h^−1^) and the highest value being for speaking during light activity (more than 1000 quanta h^−1^).

**Table 3.** Quanta emission rates (ERq) for a SARS-CoV-2 infected asymptomatic subject (cv=10^8^ copies mL^−1^) as a function of the activity level and respiratory activity.

| Activity level | Respiratory activity |  |  |  |  |
| --- | --- | --- | --- | --- | --- |
|  | Voiced counting | Whispered counting | Speaking | Breathing | Avg |
| Resting | 49.9 | 12.1 | 320 | 10.5 | 98.1 |
| Standing | 74.8 | 18.1 | 480 | 15.7 | 147 |
| Light exercise | 161 | 39.1 | $1.03 \times 10^3$ | 33.9 | 317 |

### 3.2. Results of the demonstration application

In this section, the results of the simulations performed for the microenvironments and exposure scenarios described in section 2.2 and summarized in Table 2 are reported.

#### 3.2.1. Infection risk and R_0_ for different indoor environments and exposure scenarios

As an illustrative example, Figure 2 shows the quanta concentration (*n*(t)) and infection risk (R) trends as a function of time for two different exposure scenarios simulated for the pharmacy, i.e. before lockdown (B) in natural (NV) and mechanical ventilation (MV) conditions. The trends clearly highlight that the presence of the infected individual remaining inside for 10 minutes leads to an increase in the quanta concentration in the volume: in particular, a higher peak of quanta concentration was recognized, as expected, for reduced ventilation (NV) with respect to the mechanical ventilation (MV). People entering the pharmacy after the infected individual are exposed to a certain quanta concentration during their 10-min time, and the resulting risk for their exposure (evaluated through equation (4)) is just a function of the quanta concentration trend. For example, people entering the microenvironment around the quanta concentration peak are at a higher risk than people entering the pharmacy later. Figure 2 shows an example of a customer entering at min 26 and leaving at min 36: the risk for this 10-min exposure is 2.4% in natural ventilation conditions and 1.0% in mechanical ventilation conditions. During the entire exposure time of such a scenario (3 hours and 10 minutes), 179 customers (after the infected individual) enter the pharmacy and each of them receive their own risk. In particular, the average risk of the 179 customers is 2.0% for NV conditions and 0.4% for MV conditions, then leading to a R_0_ (among the customers) of 3.52 and 0.68, to which must be added the R_0_ of the five pharmacists exposed for the entire period. Similar trends, not shown here graphically for the sake of brevity, were obtained for all the scenarios investigated, then leading to the evaluation of the R_0_ for each of them as described in the methodology section.

**Figure 2.**
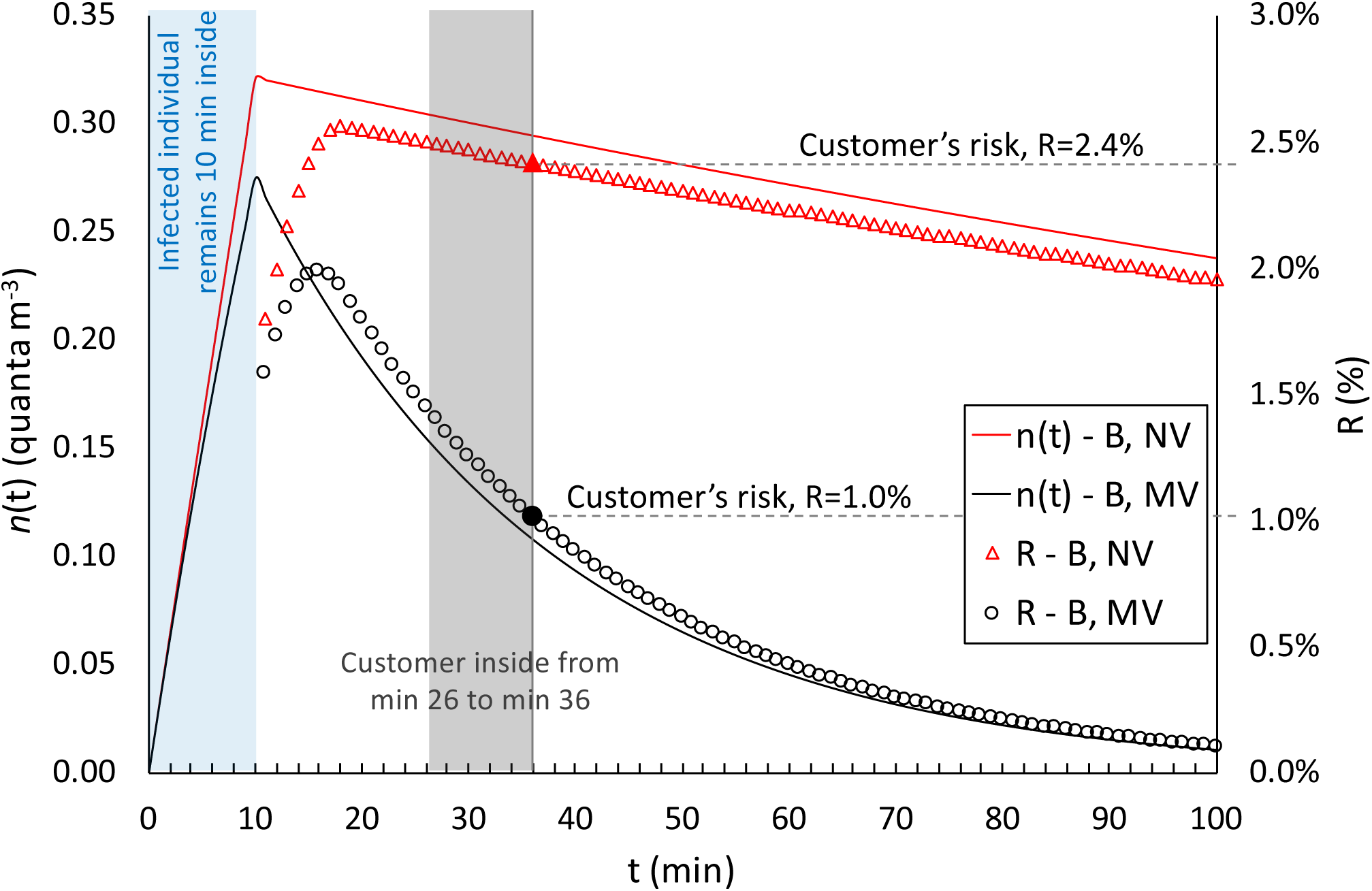
Details of application of the proposed approach in the calculation of quanta concentrations, *n*(t), and infection risks, R, in the pharmacy environment for the exposure scenarios before lockdown (B) in natural (NV) and mechanical ventilation (MV) conditions. The graph shows the entry of the infected individual (first 10 minutes) and the risk for a customer entering the microenvironment at min 26 and remaining inside for 10 minutes. The trends are shown for up to 100 minutes to highlight the peaks of the *n*(t) and R values.

Figure 3 shows the reproduction number (R_0_) data calculated for all the exposure scenarios and microenvironments under investigation (summarized in Table 2). The R_0_ data were calculated for an asymptomatic SARS-CoV-2 infected subject (c_v_=1×10^8^ copies mL^−1^) while standing; in particular, the average ER_q_ value among the different respiratory activities was considered (147 quanta h^−1^, Table **3**). The exposed subjects were also considered to be standing (IR=0.54 m^3^ h^−1^). The graph clearly highlights some critical exposure scenarios and microenvironments. Indeed, in all the microenvironments, a R_0_>1 was estimated for all the exposure scenarios before lockdown (B) when the ventilation relied only upon the building being airtight (i.e. natural ventilation conditions): R_0_ was equal to 5.55, 3.51, 59.3, 5.59, and 6.04 for pharmacy, supermarket, restaurant, post office, and bank, respectively. The huge value for the restaurant is obviously due to the simultaneous co-presence of many people (80 customers and 4 waiters) and to the long exposure time (1 h and 30 min in the current simulations). This situation is obviously improved if mechanical ventilation systems are adopted, but the R_0_ is still higher than 1 (R_0_=3.40). Similar results are obviously expected for all the indoor environments characterized by high crowding indexes and long-lasting exposures such as schools, swimming pools, gyms – venues that, in fact, were also concomitantly locked down by the government. Actually, adopting mechanical ventilation solutions that purportedly provide an adequate indoor air quality (i.e. providing AER values suggested by the standards (UNI, 1995)) did not satisfactorily reduce the R_0_ in the other microenvironments investigated. Indeed, the R_0_ values obtained from the simulations performed for the pharmacy, supermarket, post office, and bank equipped with mechanical ventilation systems in the conditions before lockdown, with mechanical ventilation in operation, were still >1.

**Figure 3.**
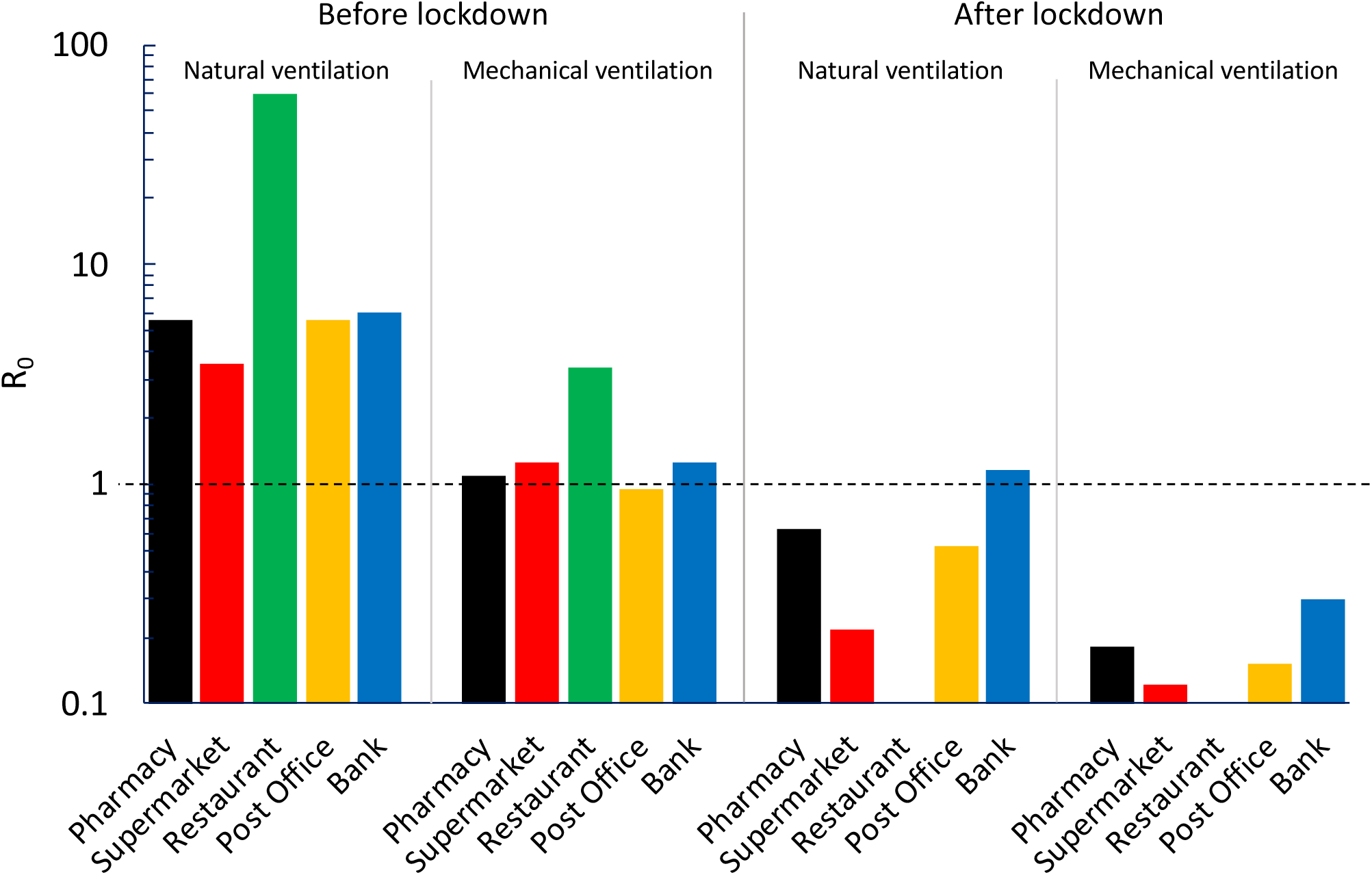
R_0_ calculated for all the exposure scenarios (natural ventilation, mechanical ventilation; before lockdown, after lockdown) and microenvironments (pharmacy, supermarket, restaurant, post office, bank) under investigation considering an asymptomatic SARS-CoV-2 infected subject (cv=1×10^8^ copies mL^−1^) while standing (IR=0.54 m^3^ h^−1^; ERq=147 quanta h^−1^) and the exposed population, also standing.

The new regulations and methods of accessing the indoor environments that were applied in the conditions after lockdown (i.e. queuing outside, limited time spent in the environments, lower crowding index) were very effective; indeed, the R_0_ values were reduced by roughly 80%–90% (for both natural and mechanical ventilation conditions) with respect to the corresponding pre-lockdown scenarios.

As an example, for the natural ventilation scenario, the only critical microenvironment was the bank, since the R_0_ was >1; this was due to a crowding index that was higher than the post-office, which had a larger floor area but same number of customers. In contrast, all the R_0_ values for indoor environments equipped with mechanical ventilation systems were much lower than 1 (0.18, 0.12, 0.15, and 0.30 for pharmacy, supermarket, post office, and bank, respectively). Therefore, if in a single day the infected individual visited different environments, the resulting R_0_ would be lower than 1 only if all the microenvironments were equipped with mechanical ventilation systems. Once again, these results highlight the importance of proper ventilation of indoor environments and are in line with the scientific literature that recognizes the importance of ventilation strategies in reducing indoor-generated pollution (Stabile et al., 2017)(Stabile et al., 2019).

The values obtained with this approach could vary significantly as a function of (i) the activity levels of both the infected subject and the exposed subjects; and (ii) the viral load in the sputum of the infected subject; therefore, in future studies, more specific exposure scenarios could be simulated on the basis of the findings proposed and discussed in this study.

## 4. Conclusions

The present study proposed the first approach aimed at filling the gap of knowledge still present in the scientific literature about evaluating the viral load emitted by infected individuals. This information could provide key information for engineers and indoor air quality experts to simulate airborne dispersion of diseases in indoor environments. To this end, we have proposed an approach to estimate the quanta emission rate (expressed in quanta h^−1^) on the basis of the emitted viral load from the mouth (expressed in RNA copies in mL^−1^), typically available from virologic analyses. Such approach also takes into account the effect of different parameters (including inhalation rate, type of respiratory activity, and activity level) on the quanta emission rate. The suitability of the findings was checked and confirmed as it was able to predict the values of quanta emission rates of previous well-known diseases in accordance with the scientific literature. The proposed approach is of great relevance as it represents an essential tool to be applied in enclosed space and it is able to support air quality experts and epidemiologists in the management of indoor environments during an epidemic just knowing its viral load, without waiting for the end of the outbreak.

For this purpose, it has been applied to the Italian case which, at the time of writing, represents the country with the highest number of deaths from SARS-CoV-2 in the world, highlighting the great importance of ventilation in indoor microenvironments to reduce the spread of the infection.

## Data Availability

No external datasets or supplementary material online are available

## References

Adams, W.C., 1993. Measurement of Breathing Rate and Volume in Routinely Performed Daily Activities. Final Report. Human Performance Laboratory, Physical Education Department, University of California, Davis. Human Performance Laboratory, Physical Education Department, University of California, Davis. Prepared for the California Air Resources Board, Contract No. A033-205, April 1993.

Chao, C.Y.H., Wan, M.P., Morawska, L., Johnson, G.R., Ristovski, Z.D., Hargreaves, M., Mengersen, K., Corbett, S., Li, Y., Xie, X., Katoshevski, D., 2009. Characterization of expiration air jets and droplet size distributions immediately at the mouth opening. Journal of Aerosol Science 40, 122–133. https://doi.org/10.1016/j.jaerosci.2008.10.003

d’Ambrosio Alfano, F.R., Dell’Isola, M., Ficco, G., Tassini, F., 2012. Experimental analysis of air tightness in Mediterranean buildings using the fan pressurization method. Building and Environment 53, 16–25. https://doi.org/10.1016/j.buildenv.2011.12.017

Duguid, J.P., 1945. The numbers and the sites of origin of the droplets expelled during expiratory activities. Edinburgh Medical Journal LII (II), 385–401.

Gammaitoni, L., Nucci, M.C., 1997. Using a mathematical model to evaluate the efficacy of TB control measures. Emerging Infectious Diseases 335–342.

Hickey, A.J., Mansour, H.M., 2019. Inhalation Aerosols: Physical and Biological Basis for Therapy, Third Edition. Taylor &Francis Ltd.

Hirose, R., Daidoji, T., Naito, Y., Watanabe, Y., Arai, Y., Oda, T., Konishi, H., Yamawaki, M., Itoh, Y., Nakaya, T., 2016. Long-term detection of seasonal influenza RNA in faeces and intestine. Clinical Microbiology and Infection 22, 813.e1-813.e7. https://doi.org/10.1016/j.cmi.2016.06.015

International Commission on Radiological Protection, 1994. Human respiratory tract model for radiological protection. A report of a Task Group of the International Commission on Radiological Protection. Annals of the ICRP 24, 1–482. https://doi.org/10.1016/0146-6453(94)90029-9

Jennison, M.W., 1942. Atomizing of mouth and nose secretions into the air as revealed by high speed photography. Aerobiology 17, 106–128.

Knibbs, L.D., Morawska, L., Bell, S.C., 2012. The risk of airborne influenza transmission in passenger cars. Epidemiology and Infection 140, 474–478. https://doi.org/10.1017/S0950268811000835

Knibbs, L.D., Morawska, L., Bell, S.C., Grzybowski, P., 2011. Room ventilation and the risk of airborne infection transmission in 3 health care settings within a large teaching hospital. American Journal of Infection Control 39, 866–872.

Morawska, L., 2006. Droplet fate in indoor environments, or can we prevent the spread of infection? Indoor Air 16, 335–347. https://doi.org/10.1111/j.1600-0668.2006.00432.x

Morawska, L., Johnson, G.R., Ristovski, Z.D., Hargreaves, M., Mengersen, K., Corbett, S., Chao, C.Y.H., Li, Y., Katoshevski, D., 2009. Size distribution and sites of origin of droplets expelled from the human respiratory tract during expiratory activities. Journal of Aerosol Science 40, 256–269. https://doi.org/10.1016/j.jaerosci.2008.11.002

Pan, Y., Zang, D., Yang, P., Poon, L.M., Wang, Q., 2020. Viral load of SARS-CoV-2 in clinical samples Yang Pan Daitao Zhang Peng Yang Leo L M Poon Quanyi Wang. The Lancet.

Papineni, R.S., Rosenthal, F.S., 1997. The size distribution of droplets in the exhaled breath of healthy human subjects. Journal of Aerosol Medicine.

Riley, C., Murphy, G., Riley, R.L., 1978. Airborne spread of measles in a suburban elementary school. American journal of epidemiology 431–432.

Rothe, C., Schunk, M., Sothmann, P., Bretzel, G., Froeschl, G., Wallrauch, C., Zimmer, T., Thiel, V., Janke, C., Guggemos, W., Seilmaier, M., Drosten, C., Vollmar, P., Zwirglmaier, K., Zange, S., Wölfel, R., Hoelscher, M., 2020. Transmission of 2019-nCoV Infection from an Asymptomatic Contact in Germany. N Engl J Med 382, 970–971. https://doi.org/10.1056/NEJMc2001468

Rothman, K.J., Greenland, S., Lash, T.L., 2008. Modern Epidemiology, 3rd ed. Lippincott Williams & Wilkins.

Rudnick, S.N., Milton, D.K., 2003. Risk of indoor airborne infection transmission estimated from carbon dioxide concentration. Indoor Air 13, 237–245. https://doi.org/10.1034/j.1600-0668.2003.00189.x

Stabile, L., Buonanno, G., Frattolillo, A., Dell’Isola, M., 2019. The effect of the ventilation retrofit in a school on CO2, airborne particles, and energy consumptions. Building and Environment 156, 1–11. https://doi.org/10.1016/j.buildenv.2019.04.001

Stabile, L., Dell’Isola, M., Russi, A., Massimo, A., Buonanno, G., 2017. The effect of natural ventilation strategy on indoor air quality in schools. Science of the Total Environment 595, 894–902. https://doi.org/10.1016/j.scitotenv.2017.02.030

Sze To, G.N., Chao, C.Y.H., 2010. Review and comparison between the Wells–Riley and dose-response approaches to risk assessment of infectious respiratory diseases. Indoor Air 20, 2–16. https://doi.org/10.1111/j.1600-0668.2009.00621.x

Tang, J.W., Noakes, C.J., Nielsen, P.V., Eames, I., Nicolle, A., Li, Y., Settles, G.S., 2011. Observing and quantifying airflows in the infection control of aerosol- and airborne-transmitted diseases: an overview of approaches. Journal of Hospital Infection 77, 213–222. https://doi.org/10.1016/j.jhin.2010.09.037

To, K.K.-W., Tsang, O.T.-Y., Leung, W.-S., Tam, A.R., Wu, T.-C., Lung, D.C., Yip, C.C.-Y., Cai, J.-P., Chan, J.M.-C., Chik, T.S.-H., Lau, D.P.-L., Choi, C.Y.-C., Chen, L.-L., Chan, W.-M., Chan, K.-H., Ip, J.D., Ng, A.C.-K., Poon, R.W.-S., Luo, C.-T., Cheng, V.C.-C., Chan, J.F.-W., Hung, I.F.-N., Chen, Z., Chen, H., Yuen, K.-Y., 2020. Temporal profiles of viral load in posterior oropharyngeal saliva samples and serum antibody responses during infection by SARS-CoV-2: an observational cohort study. The Lancet Infectious Diseases. https://doi.org/10.1016/S1473-3099(20)30196-1

UNI, 1995. UNI 10339 - Impianti aeraulici al fini di benessere. Generalità, classificazione e requisiti. Regole per la richiesta d’offerta, l’offerta, l’ordine e la fornitura.

van Doremalen, N., Bushmaker, T., Morris, D.H., Holbrook, M.G., Gamble, A., Williamson, B.N., Tamin, A., Harcourt, J.L., Thornburg, N.J., Gerber, S.I., Lloyd-Smith, J.O., de Wit, E., Munster, V.J., 2020. Aerosol and Surface Stability of SARS-CoV-2 as Compared with SARS-CoV-1. N Engl J Med. https://doi.org/10.1056/NEJMc2004973

Wagner, B.G., Coburn, B.J., Blower, S., 2009. Calculating the potential for within-flight transmission of influenza A (H1N1). BMC Medicine 7, 81. https://doi.org/10.1186/1741-7015-7-81

Wells, W.F., 1934. On airborne infection: study II. Droplets and Droplet nuclei. American Journal of Epidemiology 20, 611–618. https://doi.org/10.1093/oxfordjournals.aje.a118097

Woelfel, R., Corman, V.M., Guggemos, W., Seilmaier, M., Zange, S., Mueller, M.A., Niemeyer, D., Vollmar, P., Rothe, C., Hoelscher, M., Bleicker, T., Bruenink, S., Schneider, J., Ehmann, R., Zwirglmaier, K., Drosten, C., Wendtner, C., 2020. Clinical presentation and virological assessment of hospitalized cases of coronavirus disease 2019 in a travel-associated transmission cluster. medRxiv 2020.03.05.20030502. https://doi.org/10.1101/2020.03.05.20030502

